# Socioeconomic Disparities for Healthcare Utilization of Senior Adult Falls in Southeast Wisconsin, 2020-2022

**DOI:** 10.1101/2023.04.24.23289062

**Authors:** Ling Tong, Masoud Khani, Bradley Taylor, Kristen Osinski, Jazzmyne A. Adams, David R. Friedland, Jake Luo

## Abstract

**Background:** To examine the social determinant factors of the healthcare utilizations for senior patients with history of falls.

**Methods:** We analyzed effects of socioeconomic factors on the utilization rate of healthcare in a tertiary care center including 495,041 senior adults. We included Zip Code Tabulation Areas to measure socioeconomic factors on a community level. Cohort group comparison and multiple linear regression models evaluated the association between healthcare services utilization and age, sex, education, race, insurance type, distance, and income levels.

**Results:** Patients with a history of falls were older than those without a history of falls (79.4 ± 12.1 vs. 75.4 ± 11.6 years old), predominantly female (odds ratio [OR]: 1.26, 95% confidence interval [95% CI]: 1.24–1.28), white (OR: 1.35, 95% CI: 1.32–1.38). Patients with a fall history were predominantly retired (OR: 2.53, 95% CI: 2.49–2.58), publicly insured (OR: 2.88, 95% CI: 2.82–2.93), and more likely to require an interpreter during a hospital visit (OR: 2.40, 95% CI: 2.35–2.44). Using a geospatial analysis, healthcare utilization was higher in areas close to the care center. A regression model showed that a community’s median income, private insurance rate, and college education rate were positively associated with healthcare utilization.

**Conclusions:** Lower utilization of healthcare is associated with disadvantaged neighborhood social conditions, including under-insured status, residing far from a hospital, lower education, and lower income. We revealed the current inequities and disparities in the healthcare of senior adult fall patients in Southeast Wisconsin.

**Summary:** *What is the current understanding of this subject?:* Current healthcare outcome studies focus on risk factors of falls. However, there is a lack of studies on patient socioeconomic effects to the healthcare utilization.

*What does this report add to the literature?:* The study reveals the potential socioeconomic inequities and disparities in the healthcare adoption of the senior adult falls.

*What are the implications for public health practice?:* With the growing percentage of senior adult populations, specific strategies are needed to address the disparities to the adoption in underserved senior populations.

## Introduction

Falls are a leading cause of injury and death in older adults. [1] Over 25% of senior adults fall each year. [2] Half of senior adults over 80 had a history of falling.[3] Additionally, 25 – 30% of patients with a history of falls suffer moderate to severe injuries, limiting their ability to live independently within their community. Hospitalization is usually needed after a major injury[4]. In 2018, 2.4 million older adults were treated in emergency rooms for non-fatal fall injuries, with over 700,000 hospitalized,[5] and over 34,000 older adults died from unintentional fall injuries.[5]

The risk factor for falls has been comprehensively investigated. [7,8,9] The two major factors are person-specific and environmental factors. The latter include poor-fitting footwear, slippery floors, tripping hazards, loose rugs, a lack of stair railings, poor lighting.[10] Improving home environments can help to reduce the risk of falling. Personal factors include individual characteristics such as age, functional abilities, chronic medical conditions, and gait balance.[11] Most risk factors found in the literature were intrinsic and included a wide range of risk categories: demographic profile, lower extremity strength, vertigo and dizziness, vision, cognition deficiency, cardiovascular disease, medications, depression, gait, and balance caused by normal aging and pathological effects.[11,12] Each risk category had several risk factors that increased the likelihood of falling.

Compared with clinical risk factors, socioeconomic factors are less studied. Fall-related healthcare may be unavailable low-income seniors.[17] The seniors’ socioeconomic status can be measured by education, occupation, income, and location.[18] Lower socioeconomic status is associated to poorer health conditions, making them more likely to fall. Despite the importance of socioeconomic variables, very few investigated the association between socioeconomic factors and elderly fall risk. It is unclear how socioeconomic factors affect fall patients’ healthcare adoptions.

Many studies showed that health equity is preventing equal care for all populations. [19,20] Offering a more equitable healthcare services leads to more efficient healthcare systems overall. [21] Patients with lower socioeconomic status may receive poorer healthcare than the general population. Investigating the fall health disparities can benefit a large population, especially for low-income and socially disadvantaged population. Discovering the health equity issues and providing guidelines ensures a better health outcome and recommendations for specific populations.

To understand the health equity challenges and close the gap for senior adult patients, this study examined how socioeconomic factors affect the healthcare access for seniors over 60 years old with or without a history of falls. This study results could provide resources and data for policymakers and providers to make guidelines and address the senior adult care inequities.

## Methodology

The study was conducted in the Milwaukee metropolitan area. The population included all patients aged 60 or older who visited Froedtert Hospital and Medical College of Wisconsin between March 1, 2020, and March 1, 2022. Fall patient and non-fall patient cohorts were identified using diagnostic codes in electronic health records. Fall patients had at least one fall-related injury, and non-fall patients were not diagnosed with falls and related injuries during the study period.

We collected demographic and clinical data, including age, gender, race, insurance type, co-morbidities, and medication use. This study also collected zip-code tabulation area (ZCTA) data, including median household income, poverty rate, educational level, and unemployment rate. Multivariable logistic regression models were employed to examine the association between socioeconomic factors and healthcare utilization for fall patients compared to non-fall patients, together with determining odds ratios (ORs) and 95% confidence intervals (CIs) to quantify the strength and direction of the association. The Zip-Code area-based variables were from the United States Census Bureau’s 2018 American Community Survey. The multiple regression model was adjusted by patients based on age, gender, race, insurance type, and co-morbidities to examine potential effects.

### Data Collection

All in-person hospital visits by senior adults over 60 were included in the study. This study divided patients into two exclusive groups. Falls group patients had at least one fall-related ICD-10 diagnosis during the study period (Appendix 1). Non-fall patients had non-fall-related diagnoses in their electronic health records. We recorded age, gender, race, insurance, ethnicity, employment status, and interpreter assistance during visits for each group. The database’s payer information classified insurance status as public, private, other, or uninsured.

This study collected socioeconomic variables from eight counties—Jefferson, Kenosha, Milwaukee, Ozaukee, Racine, Walworth, Washington, and Waukesha—with 126 zip codes. Zip code tabulation area-based variables include the following: (1) whites percentage, (2) college-educated percentage, (3) public and private insurance coverage percentage, (4) median household income, (5) Area Deprivation Index, and (6) Rural-Urban Continuum Codes.

The Area Deprivation Index was based on a measure created by the Health Resources and Services Administration. It allowed for rankings of neighborhoods by socioeconomic disadvantage in a region of interest. Area Deprivation Index ranged on a scale of 0 to 100, from the most affluent to the most disadvantaged, and according to mixed factors including income, education, employment, and housing quality. ADI was used to inform socioeconomic status, health delivery, and policy conditions, especially for the most disadvantaged neighborhood groups. The Rural-Urban Continuum Codes distinguishes metropolitan counties by the population size to metropolitan areas, and non-metropolitan counties. Appendix 2 shows the categorization of Rural-Urban Continuum Codes.

This study defined a utilization rate as a predictor variable, deemed to be the number of patients with a fall history divided by the total population within each zip code block area. The utilization rate for non-fall patients was deemed to be the number of patients without a fall history divided by the total population in each zip code block area. Consequently, this study can associate the utilization rate variable with socioeconomic variables to evaluate the effect of each socioeconomic factor.

### Statistical and Geographical Analysis

All statistical analyses were performed using R programming language. Statistical tests were two-sided, and alpha was set at 0.05. This study calculated P-values using chi-square tests for categorical variables. Within table 1 of the comparison for two cohorts, this study used the odds ratio (OR) to measure the association with patient characteristics. ORs were calculated through a two-by-two contingency table. The table compared the size of the effect between the fall history group and the non-fall history group. Concerning each patient characteristic, an OR value larger than 1 indicated that patients with the corresponding characteristics were more likely to experience falls and visit the hospital, while an OR value smaller than 1 indicated that patients with the corresponding characteristics were less likely to experience falls. The 95% confidence interval demonstrated the 95% likelihood range of the OR, based upon a normal distribution. A P-value < 0.05 indicated the difference in patient characteristics between the two groups to be statistically significant.

**Table 1:**
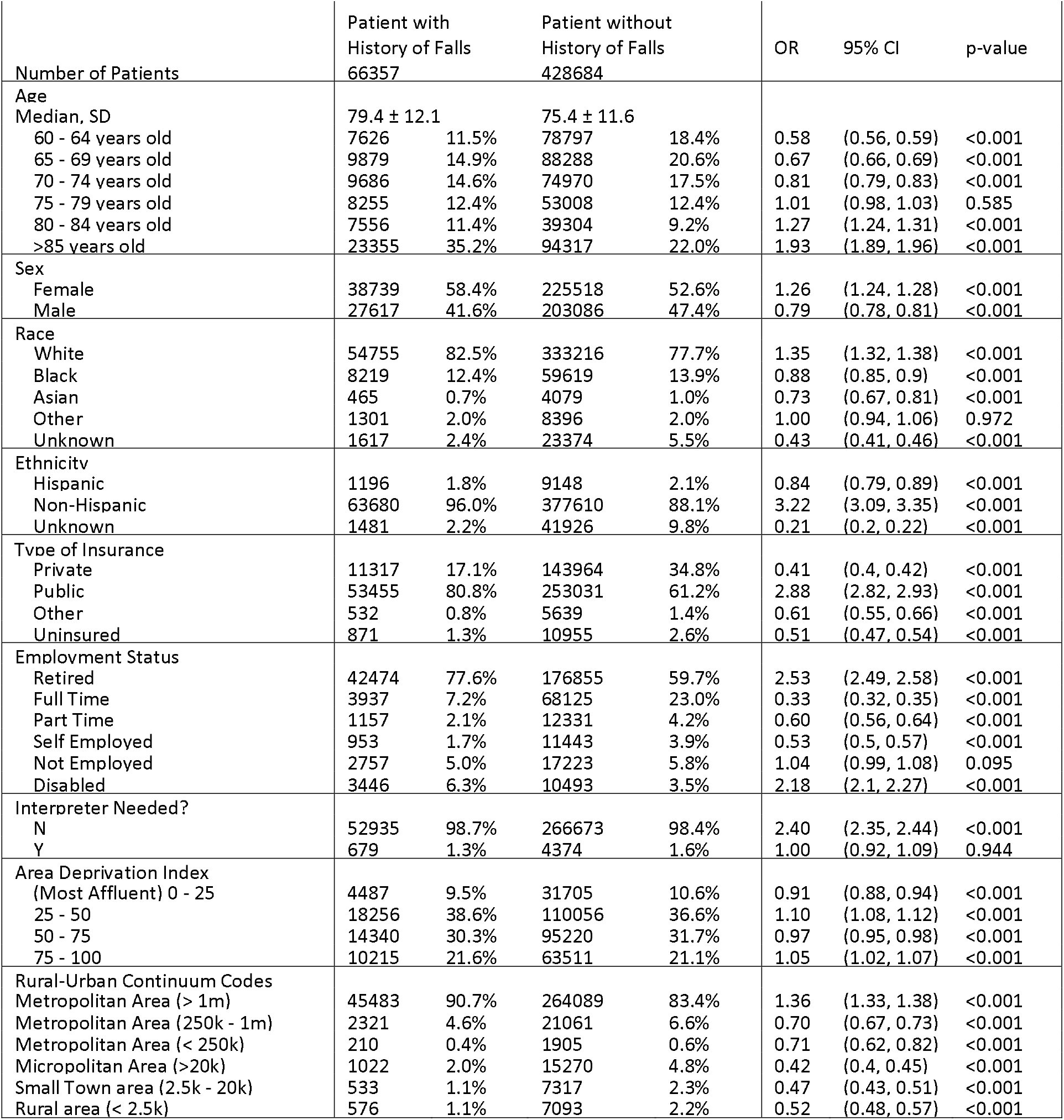
Overall patient characteristics, socioeconomic, and demographic variable comparisons for patients with or without fall history.

To further quantify the association between socioeconomic variables and utilization rates, this study completed a multiple linear regression analysis. The multiple linear regression used the utilization rate of falls and the utilization rate of other care as predictor variables, employing socioeconomic analysis as an independent variable. This study plotted scatter plots and calculated the coefficient for each socioeconomic variable. Finally, multiple regression analysis examined how social determinant factors affect healthcare utilization.

## Results

### Healthcare utilization gaps between two patient groups

Table 1 shows that 495,041 patients between March 1st, 2020, and March 1st, 2022. 66,357 (13.4%) had fall-related conditions. Patient with falls were older (79.4 ± 12.1 vs 75.4 ± 11.6 years old). Data shows falling risk increases with age. Employment variable also shows retired patients were 2.53 times more likely to experience falls and access healthcare facilities than other employed patients.

Demographic factors also affect the healthcare utilization. Females has a higher ratio of falling conditions, showing 1.26 times more than man in fall utilization. White patients used healthcare 1.35 times more than other races. Asian and black had lower utilization. Hispanics are underserved in the healthcare system because non-Hispanics were 3.22 more likely to use such services.

Economic factors also affect healthcare utilization. Public insurance patients used the services 2.88-fold more than other insurers. Uninsured patients used healthcare less frequently, with a OR value of 0.51 demonstrated that healthcare underserves uninsured patients. There was no association between Area deprivation index and the healthcare utilization.

Location also affected health care access. The hospital is in Wauwatosa, Wisconsin, a Milwaukee suburb. Over 90% of study patients live in Milwaukee. Rural patients were 1.33-fold less likely to use the services than those in metropolitan Milwaukee, indicating that rural patients may not have equal access to healthcare.

### Geographic Map-based Analysis

The utilization rate map in Figure 1 shows higher utilization around the hospital. Suburban and rural areas far from the hospital have significantly less utilizations. We further showed socioeconomic variables, including college education, white rate, income, and private insurance rate in Figure 2. From the map distribution, we found significant differences in college education rates, racial distribution, and median household income. The map analysis showed that the Northwest had a high utilization rate and private insurance rate, suggesting a correlation.

**Figure 1.**
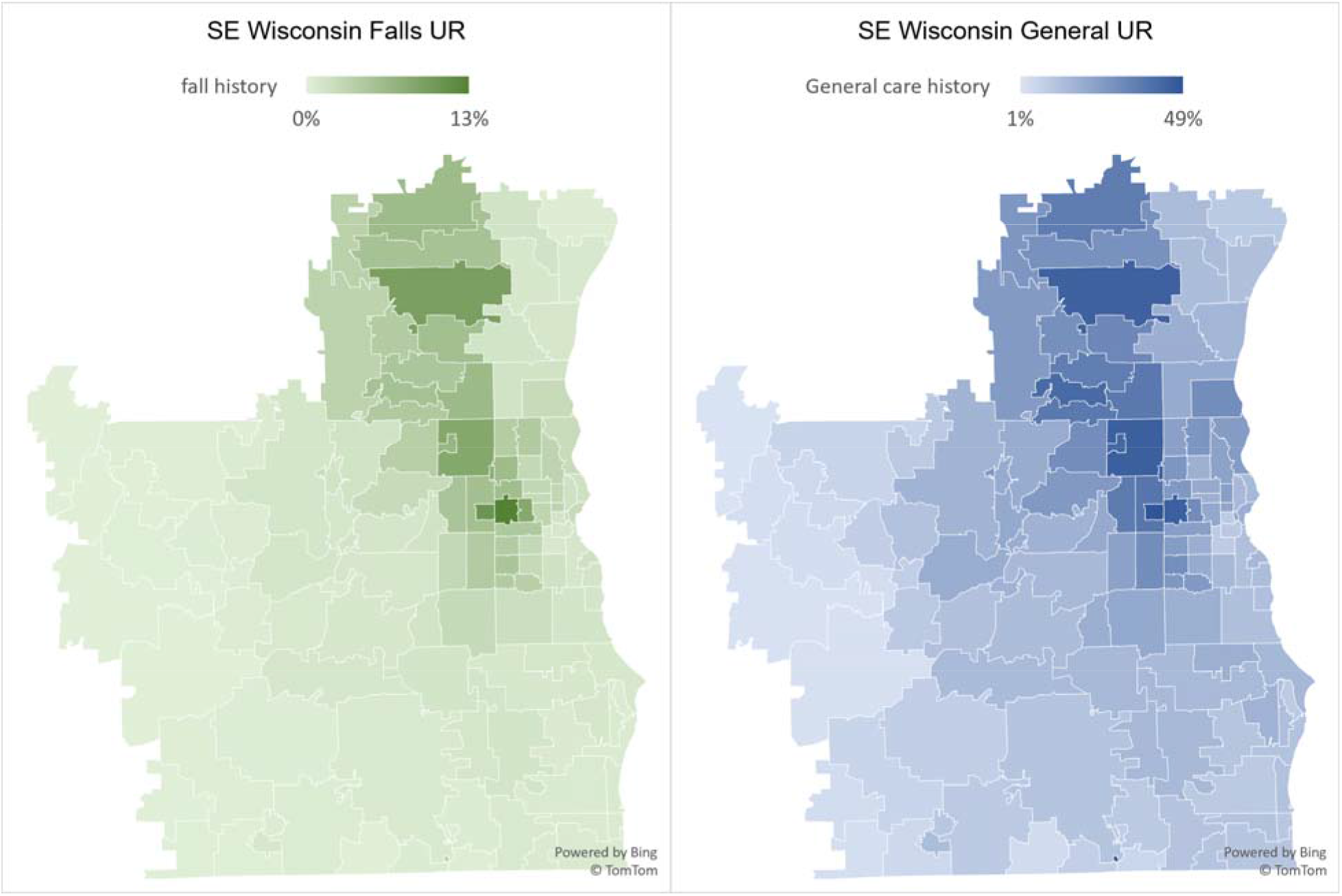
Utilization rate of general and fall-based care in Southeastern Wisconsin (SE: Southeast; UR: Utilization Rate)

**Figure 2.**
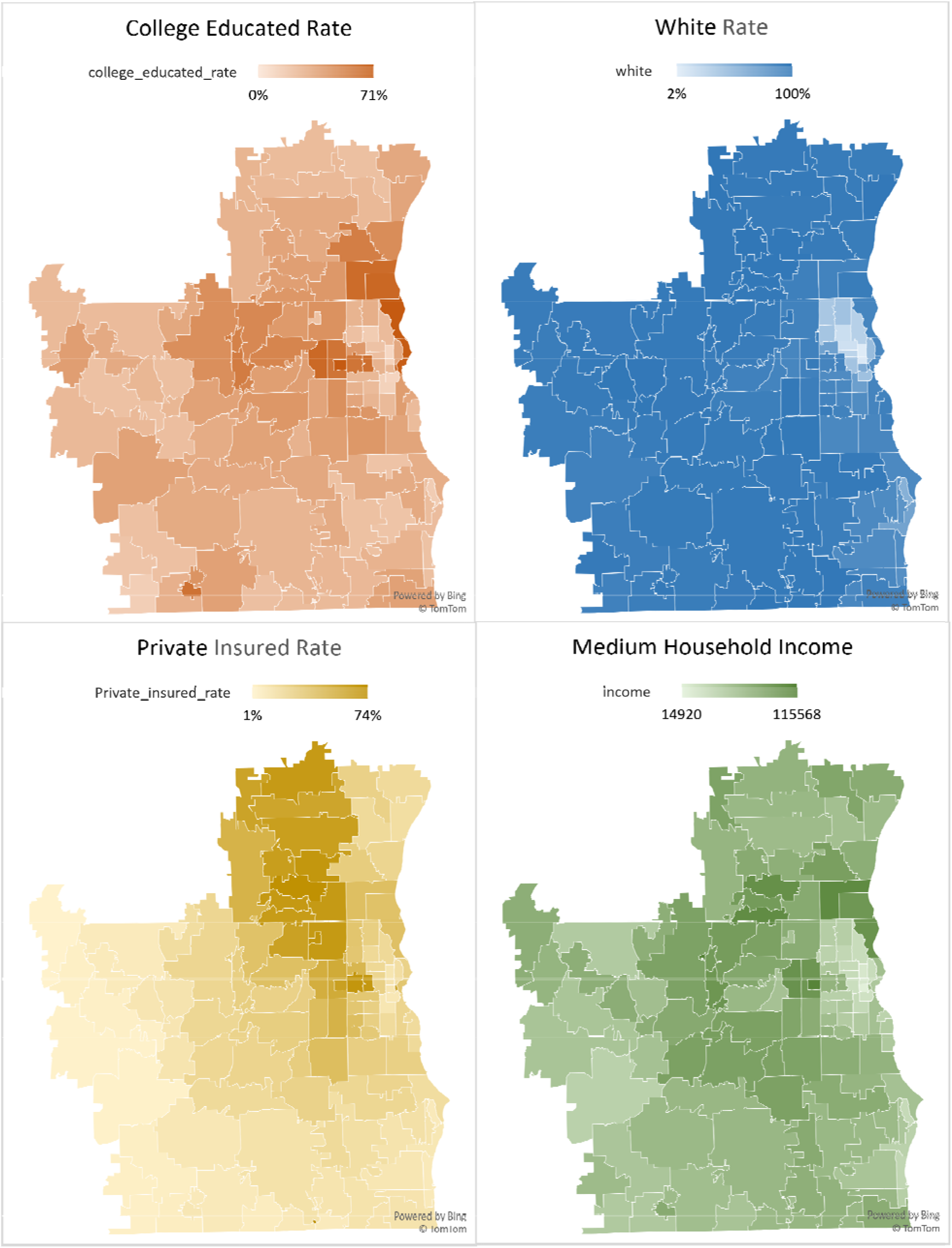
Visualization of four socioeconomic variables in Southeast Wisconsin area in a Zip code tabulated map.

### Zip Code Tabulation Area-based Analysis: Linear Regression Assessment

Figure 3 shows scatter plots and regression analyses of socioeconomic factors and fall and non-fall patient cohort utilization rates. The linear regression analysis shows private insurance rate was the most robust predictor of healthcare utilization rates for both fall-based and non-fall-based healthcare, with R-squared values of 0.68 and 0.79, respectively. College educated rate and median household income also showed a positive association with healthcare utilization rates.

**Figure 3.**
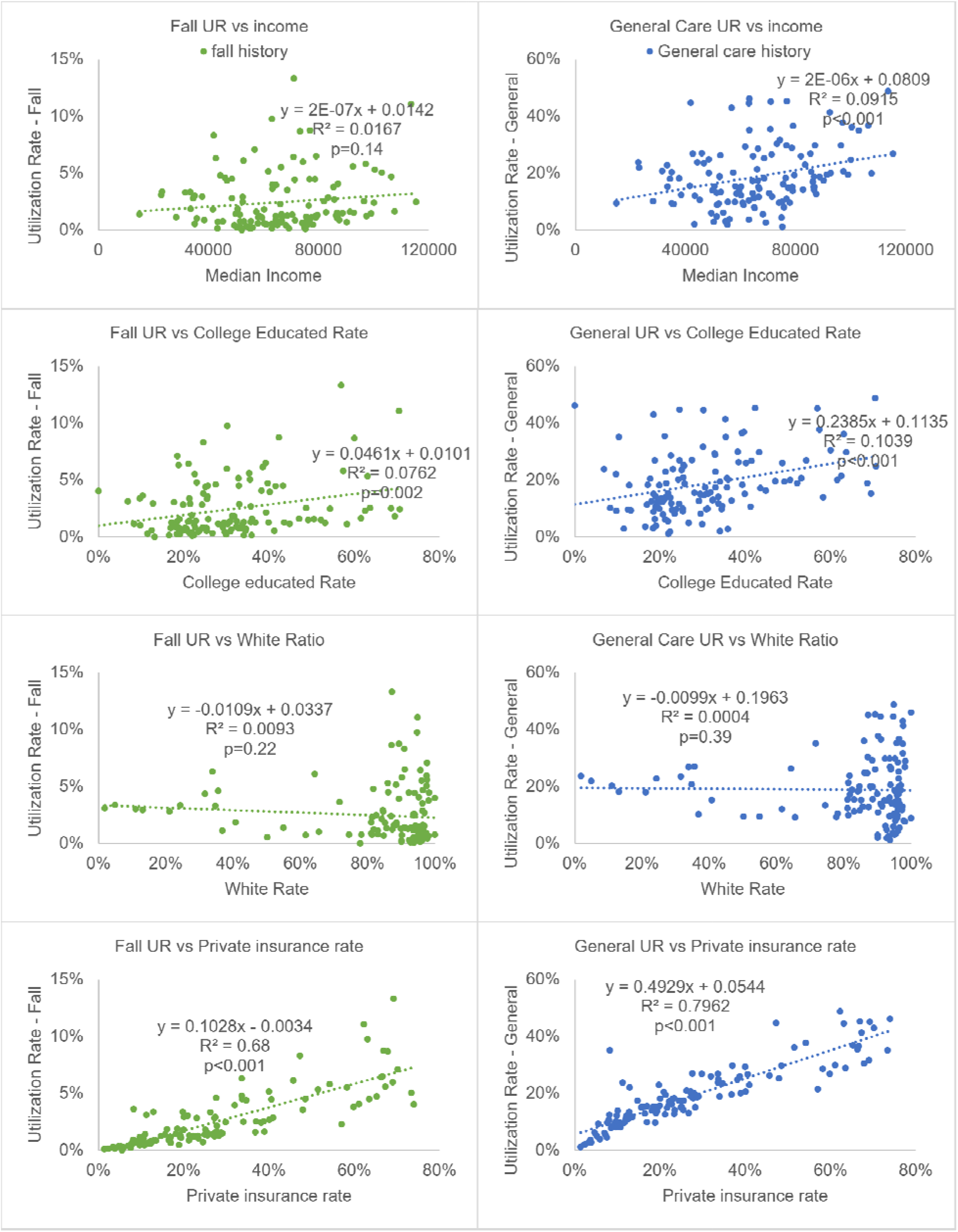
Scatter plot and regression analysis for utilization rate and socioeconomic variables across 126 zip code tabulation areas of the Southeast Wisconsin area.

In the multiple regression analysis in Table 2, the white ratio, college education rate, and private insurance rate were associated with healthcare utilization for fallers. College education and private insurance were positively associated with healthcare utilization in non-fallers. Table 2 suggests that socioeconomic status affects healthcare utilization, particularly fall-related care. Patients with private insurance and higher education levels have better access to healthcare services than those from racial and ethnic minority groups, lower income households, and uninsured individuals.

**Table 2:** Multiple regression modelling of the impact of socioeconomic variables for falls / non-fall patients within Southeastern Wisconsin area.

| Social Determinant factors | Coefficient Estimate | Standard Error | P-value |
| --- | --- | --- | --- |
| Patient of Fall History: |  |  |  |
| Income | 2.948E-07 | 9.138E-08 | 0.0016 |
| **White Ratio** | **-0.01411** | **0.006768** | **0.0492** |
| **College educated rate** | **0.01748** | **0.007898** | **0.0399** |
| **Private insurance rate** | **0.1132** | **0.006331** | **<0.0001** |
| Patients of general care |  |  |  |
| Income | 7.883E-07 | 6.525E-07 | 0.2294 |
| White Ratio | -0.1075 | 0.04836 | 0.0281 |
| **College educated Rate** | **0.1597** | **0.07073** | **0.0257** |
| **Private insurance rate** | **0.5448** | **0.04524** | **<0.0001** |

## Discussion

We examined socioeconomic factors and health utilization rates in 495,041 senior adults from a Milwaukee tertiary care center. This study showed that lower socioeconomic status is associated with lower general healthcare and fall-based care utilization. Data showed that low-income patients were underserved by healthcare. Uninsured people, low-income and low-education communities, and rural residents were included. The study confirms previous findings on socioeconomic factors and healthcare use. This study supports previous research [13, 17, 18] that older people, women, and certain racial and ethnic groups are more likely to use healthcare. This study also supports previous findings that uninsured and low-income people use healthcare less. Previous research has also examined the geographical distribution of healthcare utilization, finding that urban areas have higher utilization rates than rural areas [29]. Regression analysis showed that median household income, college education, and private insurance rates were positively correlated with healthcare utilization in each zip code tabulation area. These results indicate that economic conditions affect healthcare access and use.

The US Census Bureau reports a fast aging trend [22]. The baby boomers had 76 million US births from 1946 to 1964 [23]. Approximately 16.5% (54.1 million) of the total population reaches 65 years of age or older [24], most are at risk for falling. It is imperative to manage the risk of falls for the elderly. On the one hand, patient falls are associated with other ongoing clinical conditions, such as hypertension [25], physical or cognitive impairments [26], medication [27], and environmental hazards [28]. More clinical analysis of elderly falls may reveal key clinical risk factor correlations. Clinical factors and falls can be studied further. Utilizing these clinical factors can reduce fall risk.

Clinical factors are the focus of care [30], but equitable healthcare is neglected. Few studies have examined the relationship between fall risks and socioeconomic status, which may widen the gap between patients. According to this study, social determinants of health can affect senior adults’ healthcare access. This study focused on Southeastern Wisconsin patients’ socioeconomic disparities. Census data contains social determinant variables, making such analysis easy to replicate in other US cities and counties. This study also quantified how social gaps affect healthcare use. Close socioeconomic gaps to achieve equal healthcare, according to quantitative results.

This study found that socially marginalized senior adults underuse healthcare (Table 1). This study found that many socioeconomic determinants are linked to the healthcare utilization gap (Table 2, Figures 2-4). Healthcare professionals are needed to close the utilization gap. Equitable healthcare improves health outcomes and healthcare systems, so policies and practices must balance equity and efficiency for equal care. Socioeconomic factors are discussed below:

### Gender Differences

Female patients are 1.23-fold more likely to fall than male patients, which is consistent with other studies. These studies suggest that increased gait variability during dual-tasking may increase women’s fall risk.[31] Environmental hazards that cause falls can differ between men and women, but clinical risk factors must be identified. Gait variability increased risk in other studies [32]. Multi-tasking increases women’s gait variability, which increases their risk of falling and fractures. Other studies found that environmental hazards that cause falls in men and women vary. Men fell when their feet or chairs lost support. Berg [33] found that women tripped and fell more. No research has examined clinical factors that differ between men and women that cause such falls. Men and women may have different clinical fall factor risks. This study suggests more research on clinical factors that cause falls by gender.

### Insurance Disparities

Insurance Disparities: Public insurance patients have a 2.64-fold higher risk of falling than private insurance patients, showing how unequal healthcare access affects falls risk. The utilization rate shows that distance to the care center greatly affects healthcare access for elderly patients without private insurance, who are more likely to fall. Economic factors exacerbate healthcare disparities. The adjusted OR is higher than 2.64 because private insurers use more healthcare services, indicating that public insurance may be a risk factor for falls.

### Geographic Distribution Disparities

Patients who live in the Northeast and near the care center use more. Despite fall history, distance to the care center greatly affects utilization. The utilization rate also shows that approximately 5–10% of the total population has visited the care center for the purpose of diagnosing or treating falls, while 20–50% of the total population previously visited the care center for a non-fall-related diagnosis. Patients from outside the hospital’s neighborhood have less access. Metropolitan residents are 1.36-fold more likely to receive care.

### Statistics

This study hypothesized that economic conditions affect healthcare access and use. Privacy concerns prevented this study from using patient income data. Thus, regression analysis used zip code tabulation area factors. Median household income, college education, and private insurance rates were positively correlated with community healthcare utilization. This study concludes that economic conditions affect healthcare because they are highly correlated with patient income. College graduates earn more and understand routine care better, which leads to more hospital visits and better health outcomes.

Socioeconomic variables affected healthcare utilization in the multiple regression analysis. For example, 1% increases in college education and private insurance rates were associated with 0.017% and 0.113% increases in falls care utilization, respectively (Table 2). It is worth noting that cities have varying socioeconomic conditions and healthcare services, so our interpretations cannot apply to all areas. However, this method can be used in other cities or states to examine healthcare service utilization and socioeconomic variables and the social determinants of health.

This study showed how socioeconomic factors and social determinant factors are associated with healthcare adoption and inequalities. This analysis can reveal how social and economic factors affect healthcare visits for a patient cohort in a specific area. This analysis suggests that economic factors have a significant impact on access to healthcare and falls care, and that policy should facilitate more equal care for all patients, regardless of social position.

### Limitations

Several limitations should be considered. As with many social determinant studies, patient income was not collected, and the median income in each zip code was used as a proxy, which may introduce potential bias. Secondly, this study includes patient from regional hospital system in Milwaukee with one of the highest racial segregation scores in the US [34]. Therefore, the patient characteristics may not apply to other healthcare systems and areas. Thirdly, this study only examined a few key social determinant factors. Other potential factors, such as environmental and cultural factors, that could be considered in future studies. Fourthly, the COVID-19 pandemic may have a significant effect on different types of health services. Other health services such as cancer treatment, chronic condition management, laboratory services, and pharmacy services were not included in this study, which could potentially distort results. Finally, this study did not include clinical conditions or issues, suggesting that older patients may have used more healthcare services simply since they had a higher frequency of diagnosis and required additional services. Future studies need to investigate how to analyze and integrate cultural-based variables to achieve equal access to healthcare services from a variety of perspectives.

## Data Availability

All data produced in the present study are available upon reasonable request to the authors, and subject to the approval of Clinical Translational Science Institute in Medical College of Wisconsin.

## Conclusion

This study examined socioeconomic factors in senior adults’ falls diagnosis healthcare utilization. Fallers were more likely to have clinical conditions and socioeconomic factors, according to this study. Gender, race, income, education, location, and insurance type affect senior adults’ fall healthcare use. Further investigation is necessary to determine the causal relationships between these factors and the risk of falling. While the elderly population grows, it is imperative to examine the collective impact of potential socioeconomic variables to ensure equitable healthcare for all individuals.

## Acknowledgement

Conflict of interest: All authors declare that there is no conflict of interest.

