## Appendix 1 for "Socioeconomic Disparities for Healthcare Utilization of Senior Adult Falls in Southeast Wisconsin, 2020-2022"

**Appendices**

1) List of ICD-10 codes for the Falls Group

| ICD-10 Code | Description |
| --- | --- |
| W00-W19 | Slipping, tripping, stumbling and falls |
| Z91.81 | History of falling |
| 781.2 | (ICD-9) Abnormality of gait |
| R26.89 | Other abnormalities of gait and mobility |
| R26.81 | Unsteadiness on feet |
| R26.9 | Unspecified abnormalities of gait and mobility |
