## Appendix 2 for "Socioeconomic Disparities for Healthcare Utilization of Senior Adult Falls in Southeast Wisconsin, 2020-2022"

1) Categorization of Rural-Urban Continuum Codes

| **Metropolitan Counties*** | | Patient with Fall history | | Patient with Fall history | |
| --- | --- | --- | --- | --- | --- |
|  | Number of patients | 66357 | | 428684 | |
| **Metropolitan Counties** | |  |  |  |  |
| Code | Description |  |  |  |  |
| 1 | 1m population or more | 45483 | 94.7% | 264089 | 92.0% |
| 2 | 250k to 1m population | 2321 | 4.8% | 21061 | 7.3% |
| 3 | fewer than 250k population | 210 | 0.4% | 1905 | 0.7% |
| **Nonmetropolitan Counties** | |  |  |  |  |
| Code | Description |  |  |  |  |
| 4 | 20k or more, adjacent to a metro area | 878 | 52.9% | 12356 | 51.4% |
| 5 | 20k or more, not adjacent to a metro area | 144 | 8.7% | 2914 | 12.1% |
| 6 | 2.5k to 20k, adjacent to a metro area | 59 | 3.6% | 1086 | 4.5% |
| 7 | 2.5k to 20k, not adjacent to a metro area | 474 | 28.6% | 6231 | 25.9% |
| 8 | Less than 2.5k, adjacent to a metro area | 76 | 4.6% | 1010 | 4.2% |
| 9 | Less than 2.5k, not adjacent to a metro area | 28 | 1.7% | 423 | 1.8% |
