## Appendix 3 for "Socioeconomic Disparities for Healthcare Utilization of Senior Adult Falls in Southeast Wisconsin, 2020-2022"

3. Quartile categorization of Area Deprivation Index

| **Area Deprivation Index** | | Patient with Fall history (%) | | Patient without Fall history (%) | |
| --- | --- | --- | --- | --- | --- |
|  | (Most Affluent) 0 - 25 | 4487 | 9.5% | 31705 | 10.6% |
|  | 25 - 50 | 18256 | 38.6% | 110056 | 36.6% |
|  | 50 - 75 | 14340 | 30.3% | 95220 | 31.7% |
|  | (Most Deprived) 75 - 100 | 10215 | 21.6% | 63511 | 21.1% |
